# Retinol Depletion in Severe COVID-19

**DOI:** 10.1101/2021.01.30.21250844

**Authors:** Aziz Rodan Sarohan, Hakan Akelma, Eşref Araç, Özgür Aslan

## Abstract

**Background and Purpose:** Vitamin A is depleted during infections. Vitamin A has been used successfully in measles, RSV and AIDS patients and is an effective vaccine adjuvant. In this study, low retinol levels were found in patients with severe COVID-19. Retinoid signaling impairment in COVID-19 disrupts Type-I interferon synthesis.

**Material and Method:** Two groups were formed in the study. The patient group consisted of 27 (Group 1) severe COVID-19 patients hospitalized in the intensive care unit with respiratory failure, and the control group consisted of 23 (Group 2) patients without COVID-19 symptoms. Serum retinol levels were analyzed by ELIZA and HPLC in both groups.

**Findings:** Retinol levels were found to be significantly lower in the patient group (P <0.001). There was no difference in retinol between two different age groups in the patient group (P> 0.05). There was no significant difference in retinol between men and women (P> 0.05). Comorbidity did not affect serum retinol levels (P >0.05).

**Conclusion:** Serum retinol levels were low in patients with severe COVID-19. Drugs preventing retinol excretion were not stopped in the patient group. Some patients took vitamin A externally. Despite this, retinol was low in COVID-19 patients. Retinol depletion impairs Type-I interferon synthesis by impairing retinoid signaling. Retinoid signaling may be the main pathogenetic disorder in COVID-19. This pathogenesis can serve as a guide for adjuvants, drug targets, and candidate drugs. Retinol, retinoic acid derivatives, and some CYP450 inhibitors may work on COVID-19.

## Introduction

In this study, serum retinol levels were found to be low in COVID-19 patients. The metabolic dynamics of retinol and retinoic acids with the CYP450 enzyme system and their interactions with the immune system in COVID-19 have not yet been studied. Retinol depletion and retinoid signaling in COVID-19 cause the immune system to collapse, particularly type I interferon synthesis. Similarly, retinol depletion and retinoid signal impairment cause the destructive inflammatory response of the adaptive immune system. Here, we draw attention to the role of retinoid signaling disorder that develops as a result of retinol depletion in the pathogenesis of COVID-19. Retinol depletion and retinoid signaling mechanism offer a new and different perspective on COVID-19 treatment and prophylaxis.

COVID-19, which emerged in December 2019 and was caused by SARS-CoV-2, was declared an epidemic by the World Health Organization (WHO) in March 2020 [1, 3]. More than one and a half million people died in the epidemic, and millions of people also became infected. No specific treatment has yet been found for COVID-19, which causes serious socioeconomic consequences [1, 2]. The COVID-19 pandemic continues to be a major problem worldwide [3]. Reduce high death and disease rates in the largest epidemic, COVID-19. The search for drugs continues throughout the world. As new drug development is a long process, the idea of repositioning existing drugs and vaccination studies have come to the fore[4].

The protective effects of Vitamin A against infections have been known for a long time. Vitamin A deficiency leads to a tendency to infections as well as increases the clinical severity of diseases. The World Health Organization (WHO) added vitamin A prophylactic to its pandemic prevention programs in the 1950s, and achieved successful results in the measles pandemic. Vitamin A reduced mortality rates due to pneumonia by 50% in the measles pandemic [5, 6, 7]. Vitamin A deficiency is also common in HIV infection. A reduction in helper T lymphocyte, which is distinctive for HIV, was associated with Vitamin A deficiency [8]. Retinol has been used successfully in AIDS patients. Vitamin A replacement was also effective for other viral infections in AIDS [8].

Vitamin A deficiency reduces host resistance to viral infections and vitamin A levels decrease during viral infections [6, 7, 9]. In Vitamin A deficiency, the frequency of Measles, Varicella, RSV, AIDS and pneumonia as well as the mortality rates due to these diseases are increased. Besides, vitamin A stores may become empty during infection. Previous studies found that retinoic acid levels were excessively lowered during measles, RSV, HIV and pneumonia [6, 7]. During systemic infections, high fever increases metabolic use and urinary excretion and reduces apparent retinol stores [7, 9]. Measles are disruptive especially for Vitamin A metabolism, affecting negatively the use as well as storage of Vitamin A [6, 9, 10]. This creates a bad vicious circle for infections [11, 12, 13].Serum retinol levels are reduced only after Vitamin A deficiency progresses because the liver contains big Vitamin A stores. When serum retinol levels were found low, this means that retinol stores in the liver were significantly depleted [12,13].

Vitamin A deficiency is associated with immune system deficiency. Vitamin A deficiency causes disruption of mucosal barriers in the gastrointestinal and respiratory systems [9]. In addition, there is a decrease in the function of monocytes, macrophages, and natural killer cells, and a decrease in the number and function of T helper and B lymphocyte cells [14,16]. Effective antibody response is also impaired by the number and dysfunction of antigen presenting T lymphocytes, B lymphocytes, and plasma cells [15, 16]. All of these changes in the immune system functions due to Vitamin A deficiency are also seen in COVID-19 patients [17]. In severe COVID-19, neutrophil and white blood cells are elevated, while total lymphocyte count, CD4 + T cells, CD + 8 T cells, regulatory T cells (T reg), memory T cells, natural killer and B cells are lower, as well as, antibody synthesis and thus humoral immune system are also impaired [17].

Retinoic acids, a vitamin A derivative, have a central and indispensable role in immune defense mechanism. Retinol derivatives have a central regulation on many organ systems, especially the immune system, by the retinoid signaling mechanism. Vitamin A primarily strengthens the cellular and humoral immune system. Vitamin A provides mucosal immunity and mechanical protection by creating a secretory IgA and a mucosal barrier [14, 16]. Vitamin A and its derivatives show their protective effects against infections mainly through Type-I interferon synthesis [18,19]. Retinoic acids, which are derivatives of vitamin A, participate in the synthesis of Type-I interferon (IFNα and IFNβ), which are the strongest endogenous mediators in the immune defense mechanism [18, 19, 20].

Type-I interferon is synthesized via nuclear retinoic acid receptors (RXR, RAR) and transcription factors in the RIG-I pathway due to retinoic acids [18,19]. As the name suggests, RIG-I is the main cytosolic recognition receptor expressed in relation to retinoic acids and recognizing RNA viruses [18]. Type-I IFN is the most potent endogenous antiviral agent of the host, providing clearance of the pathogen from the body and the development of a sustained immune response [20]. Type-I IFN prevents viral replication through regulation of the host immune system and activation of cytotoxic T cell, induces antibody synthesis by enhancing B lymphocyte and plasma cells through T helper cells and antigen presenting cells (APC) [19,20].

## Material and Method

In the study, two groups were formed. 27 severe COVID-19 patients who were hospitalized in the intensive care unit, and had a poor general condition and respiratory failure were included in the patient group (G1, N:27), 23 individuals who were admitted to polyclinic without any COVID-19 related symptom and clinical symptom were included in the control group (G2, N:23). The control group was not subjected to COVID-19 RT-PCR test. Serum retinol levels were checked in both groups.

The venous blood samples taken from the patients were transferred to light-proof tubes. The tubes were covered with protective aluminum foil to protect Vitamin A from light. The blood samples were centrifuged at 1000 g after waiting for 30 minutes. The serum samples were kept at −80LJC in the waiting period. Retinol in well-soluble serum samples was analyzed using commercially available enzyme-linked immunosorbent assay (ELISA) test kit and the High-Performance Liquid Chromatography method (Agilent 1200 Series HPLC System, USCN Life Science, Hankou, Wuhan, China).The unit for Vitamin A was determined as mg/L.

In the patient group (N=27), the drugs containing the active ingredient Favipiravir and Hydroxychloroquine that were used for the treatment of COVID-19 were not discontinued (due to ethical concerns). Favipiravir and Hydroxychloroquine are the drugs that inhibit cytochrome oxidase P450 enzymes in the liver and prevent the excretion of retinol via the liver. Although these drugs were used in the patient group, retinol levels were found significantly low (P<0.05).

The diet regimens of the patients were not intervened in any way during the study period. The Vitamin A and polyunsaturated fatty acid contents of the nutrition solutions administered to the patients were determined retrospectively. Any diet regimen with Vitamin A supplement or Vitamin A restriction was not applied to those in the patient group who were conscious and could be fed orally. These patients continued to eat regular hospital meals. However, 12 patients in the patient group, who had difficulty to eat orally or could not be fed orally due to intubation, were fed through a nasogastric tube and parenteral route. Those who were fed through nasogastric tube received polyunsaturated fatty acid (omega3) in formulas. Those who were fed through the parenteral route received retinol and polyunsaturated fatty acid (omega3) in total parenteral nutrition (TPN) solution. The retinol and polyunsaturated fatty acid contents of the nutrition solutions administered to the patients were determined by retrospective evaluation and evaluated statistically.

### Statistical Analysis

The data obtained in the study were statistically analyzed using IBM SPSS 22.00 for Windows program (Statistical Package for Social Sciences, Chicago, IL, USA). The Shapiro-Wilk Test was used for the normal distribution of the data. It was found that age distribution and retinol measurements were not compliant with the normality assumption for the patient group. The Mann-Whitney U Test was performed for the comparisons between the sub-groups of the patient group that was not compliant with the normality assumption. When P<0.05, the result was considered to be statistically significant.

### Findings

A total of 50 individuals were included in the study. 27 severe COVID-19 patients who were hospitalized in the intensive care unit and had respiratory failure and poor general condition were included in the patient group, while 23 individuals who were admitted to the polyclinic and had no COVID-19 symptoms were included in the control group. Those in the control group were not subjected to COVID-19 RT-PCR test. Serum retinol levels were checked in both groups. The patient group consisted of a total of 27 (G1:27) individuals (14 female, 13 male). The control group consisted of a total of 23 (G2:23) individuals (15 female, 8 male). 18 individuals in the patient group had a comorbid disease that posed a high risk for mortality and morbidity in COVID-19. All patients in the patient group received Hydroxychloroquine and Favipiravir, both of which inhibit CYP450 enzymes. Ten people in the patient group took vitamin A and unsaturated fatty acids in dietary supplements. The post-study evaluation revealed that 12 people in the patient group had died.

### Retinol

A statistically significant difference was found in serum retinol levels between the two groups (P<0.001). The average retinol level was found as 0.37 mg/L in the patient group and 0.52 mg/L in the control group (Table 1), (Figure 1).

**Table 1:**
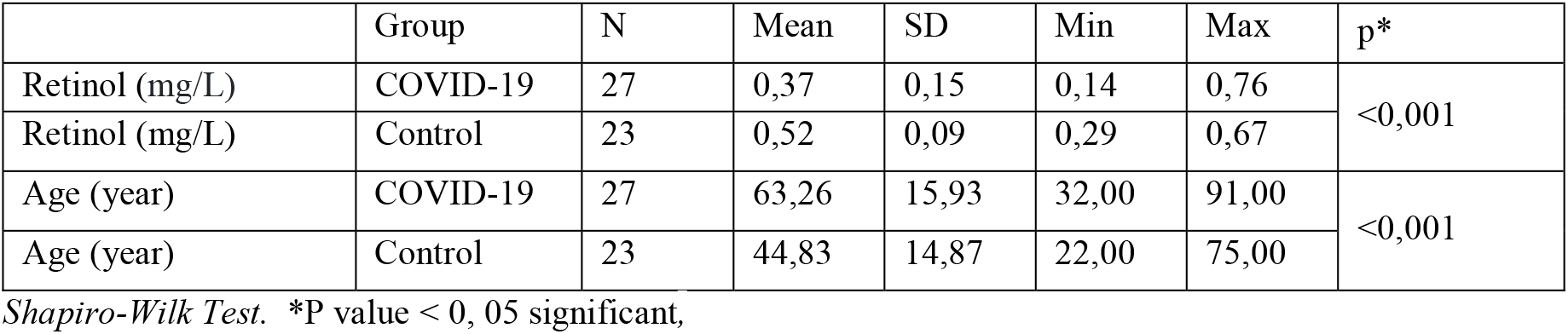
There is a significant difference in both retinol and age between Group 1 and Group 2 (P<0.001).

|  | Group | N | Mean | SD | Min | Max | p* |
| --- | --- | --- | --- | --- | --- | --- | --- |
| Retinol (mg/L) | COVID-19 | 27 | 0,37 | 0,15 | 0,14 | 0,76 | <0,001 |
| Retinol (mg/L) | Control | 23 | 0,52 | 0,09 | 0,29 | 0,67 |  |
| Age (year) | COVID-19 | 27 | 63,26 | 15,93 | 32,00 | 91,00 | <0,001 |
| Age (year) | Control | 23 | 44,83 | 14,87 | 22,00 | 75,00 |  |

*Shapiro-Wilk Test.* \*P value < 0, 05 significant,

**Figure 1:**
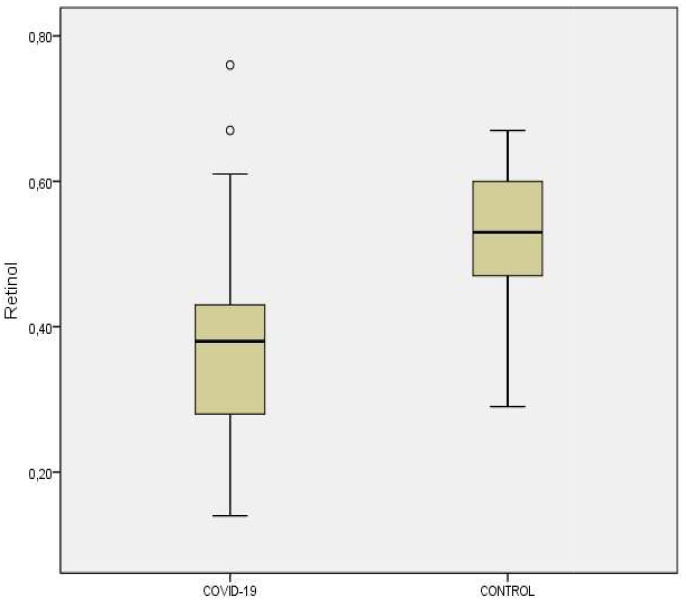
There is a significant difference in retinol levels between group1 and group2 (P<0.001).

### Age

A significant difference was found in age between the two groups. The average age of the patient group was 63.2 years, while the average age of the control group was 44.8 years.

The patient group (N=27) was divided into two age groups to correct the age-related variability of retinol levels. Under 60 years (N=10) and over 60 years (N=17) old. There was no significant difference in retinol levels between these two subgroups of the patient group (P> 0.05), (Table 2), (Figure 2).

**Table 2:** There was no significant difference in retinol levels between the two subgroups in the patient group (P> 0.05).

|  |  |  |
| --- | --- | --- |
| Retinol | ≤60 year (N= 10) | >60 year(N= 17) |
| Mean±SD | 0.38±0.21 | 0.36±0.12 |
| Median | 0.40 | 0.37 |
| Min –Max | 0.14 - 0.76 | 0.15- 0.67 |

*Mann-Whitney U Test.* P >0.05

**Figure 2:**
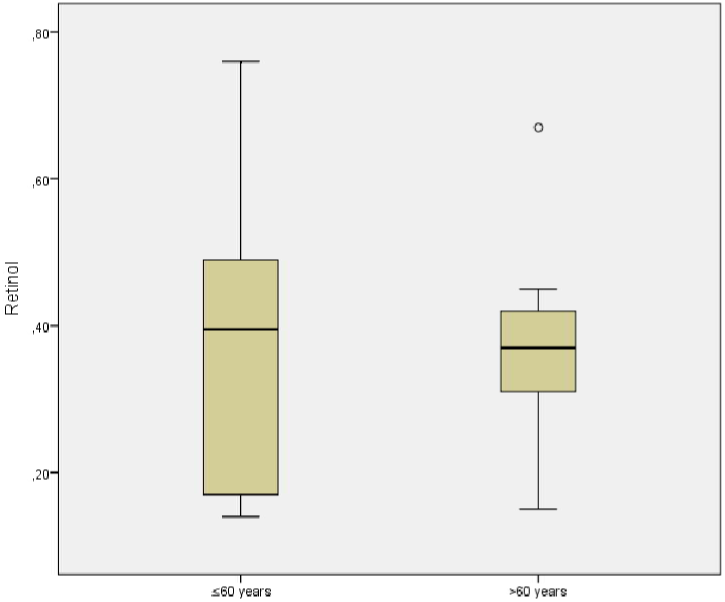
There was no significant difference in retinol levels between the two subgroups of the patient group (P>0.05).

## Results of the patient group

### Gender

In the patient group (N=27), there were 13 females and 14 males. No significant difference was found in retinol levels between the females and the males (P >0.05).

### Drug Use

In the patient group (N=27), 25 patients received Favipiravir, and 2 patients received Hydroxychloroquine. Both the drugs prevent the excretion of retinol by CYP450 inhibition. All patients in the patient group that they were taking these drugs. Although these drugs were used, retinol levels were significantly low in the patient group (P< 0.001).

### Comorbid Disease

Of 27 individuals in the patient group, 18 had a comorbid disease, while 9 had no comorbid disease. No difference was found in serum retinol levels between the two groups (P>0.05). Only all patients (8 people) who died in the Comorbid group received vitamin A supplements from outside. The reason why there was no difference in retinol between the two groups might be the use of external Vitamin A and fatty acid derivatives.

### Vitamin A Use

Of 27 patients in the patient group, 10 received Vitamin A and unsaturated fatty acid, while 17 did not receive them. The average retinol level was found higher in the group that received Vitamin A, but there was no significant difference between them (P>0.05). Of 10 patients who received Vitamin A, 9 (90%) died, 8 of these 9 patients had also a comorbid disease. Of 17 patients who did not receive Vitamin A, 4 (24%) died. There was a significant correlation between Vitamin A use and died rates between the group that received Vitamin A and the group that did not receive Vitamin A (P<0.001). 90% of the patients who received Vitamin A supplement died, however, almost all of them (8 patients) had a comorbid disease.

### Exitus

Of 27 individuals in the patient group, 12 died and 15 were discharged. There was no significant difference in retinol levels between those who died and those who were discharged (P>0.05). Of 12 individuals who died, 10 received Vitamin A with dietary supplements, and 8 of these 10 individuals had a comorbid disease.

### Ferritin Levels and Lymphocyte Counts

In the study, serum ferritin levels and lymphocyte counts were also evaluated in the patient group. In the patient group, ferritin levels were found high and lymphocyte counts were found low. These findings were compliant with the findings of clinical studies in the literature and were associated with poor prognosis in COVID-19.

However, since these two parameters were not in the scope and purpose of the study and had no direct effects on serum retinol levels, they were not included in the statistical calculations.

## Discussion

Endogenous regulation of retinoic acids is provided through the CYP450 enzyme system. With the inhibition of the P450 system, the endogenous levels of retinoic acids increase, resulting in physiological outcomes and therapeutic efficacy. Inhibition of CYP26 sub gruop enzymes especially prevents retinoid metabolism [21, 22]. Serum endogenous retinoic acid levels may be elevated in patients starting COVID-19 treatment in the early period due to CYP450 inhibition due to drugs used. Therefore, patients taking CYP450 inhibitor drugs in the early stage of the disease were not included in the clinical study. Severe COVID-19 patients who were hospitalized in the intensive care unit, whose general condition was poor and who needed respiratory support were included in the study.

CYP450 inhibitory drugs (Favipiravir and HCQ) were not discontinued in the patient group due to ethical concerns. Another factor that may affect serum retinol levels in the patient group is the use of vitamin A and long-chain fatty acids in combination with dietary supplements. In the patient group, these CYP450 inhibiting drugs and Vitamin A in dietary supplements were not intervened. Despite these two factors, retinol levels were found significantly lower in the patient group than the control group (p< 0.05). These results show that retinol was consumed and liver retinol stores are depleted in patients with severe COVID-19.

### Age

A significant difference was found in age between the two groups. The average age of the patient group was 63.2 years, while the average age of the control group was 44.8 years. The patient group was divided into two age groups to correct the age-related variability of retinol levels. Over 60 years old and under 60 years old. No significant difference was found in retinol levels between these two sub-groups that were divided by age criteria (P>0.05). This result shows that the retinol difference between the patient group and the control group is not related to age and that this significant decrease in retinol levels may be caused by serious disease.

A study conducted in Brazil found that vitamin A deficiency is more common with increasing age, independent of diet [23]. Despite varying levels of vitamin A deficiency among the elderly population living in major metropolitan areas in China, marginal vitamin A deficiency has also been found to be fairly common. Here, vitamin A deficiency has been found particularly common in metropolitan residents and the elderly. These findings explain why COVID-19 is severe and fatal, especially in metropolitan cities and the elderly population. Meanwhile, COVID-19 is known to have a fatal course in those living in metropolitan areas and especially in elderly patients [24]. Vitamin A deficiency reduces the host’s resistance to viral infections, and vitamin A levels drop during viral infections. This is exactly what happens with elderly patients. That’s why they largely succumb to the fight against COVID-19.

### Gender

There were 13 women and 14 men in the patient group. There was no significant difference in retinol levels between the two groups in terms of gender (P> 0.05). We think that retinoic acid excretion is lower in women than in men due to estradiol effect. Estradiol inhibits the CYP450 system more effectively and more widely than testosterone. Estradiol inhibits CYP3A4, CYP2A6 and CYP1A2, while testosterone inhibits only CYP2D6 [25]. This difference in CYP450 enzyme inhibition makes women more resistant to COVID-19 than men. This explains why COVID-19 is more severe in men (if we quit smoking). The CYP450 system is less suppressed in men than in women. Because of this dominant role of estradiol on CYP450 enzymes, we expected higher retinol levels in women than men in the study. The results may have been affected by the low number of cases, the in homogeneity of the patient group, the use of CYP450 inhibitory drugs, and the administration of dietary supplements containing vitamin A to the patients. A study with a larger and homogeneous group will yield more reliable results about the difference in retinol metabolism between men and women.

### Comorbid Disease

18 individuals in the patient group had at least one comorbid disease with a high risk of mortality and morbidity in COVID-19. Nine patients in this group had no comorbid diseases. There was no significant difference in serum retinol levels between the two groups (P >0.05). We expected low vitamin A levels in the comorbid group due to the inflammatory processes of chronic diseases. Some comorbid patients taking external vitamin A and fatty acid derivatives may have caused this result. In the comorbid group, 10 people were taking vitamin A and 8 people died. All of the patients in the discharge group (8 patients) were receiving external vitamin A supplements and all had comorbidities. More than half of the individuals in the comorbid group (10 people) used external vitamin A supplements and long-chain unsaturated fatty acids. This situation may have affected the results.

Another important factor that increases mortality with advanced age in COVID-19 is the presence of comorbid disease in the patient. As with infections, vitamin A deficiency predisposes to chronic inflammatory diseases [26]. In addition, during the course of these diseases, vitamin A is constantly used to suppress inflammatory processes. Vitamin A and retinoic acids show anti-inflammatory activity by preventing degradation and NFkB activation in the ubiquitin proteosome system [29, 47]. Therefore, vitamin A levels must be low in comorbid patients before COVID-19. Because of low retinol levels as in elderly patients, comorbid patients also cannot have a strong immune response against SARS-CoV-2.

### Exitus

Twelve of the 27 people in the patient group died. 15 people were discharged. There was no significant difference in retinol levels between those who died and those who were discharged (P> 0.05). However, the majority of the deceased group used vitamin A and long-chain fatty acids at levels that would affect retinol levels via a nasogastric tube or parenterally. We expected low retinol levels in the Exitus group. These external supplements increased retinol levels in the output group, but did not have a positive effect on survival. This may have been caused by using low doses of vitamin A. Because low flat retinol applied under inflammatory conditions is useless. Retinol should be kept within normal physiological limits or therapeutic ranges in order to have anti-infective and anti-inflammatory activity. Studies show that the effect of vitamin A use is dose dependent and that higher doses should be used to benefit in infections [27].

### Drug Use

In the patient group, 25 of the patients who received CYP450 inhibitory drugs received Favipiravir and 2 took Hydroxychloroquine. Therefore, it was assumed that all patients in the patient group were taking CYP450 inhibitory drugs (which inhibit retinol excretion). Despite this, retinol levels were found to be significantly lower in the patient group compared to the control group (P <0.001). Most of the drugs currently used on SARS-CoV-2 in the treatment of COVID-19 do not have an antiviral effect. Most of them act by inhibiting the CYP450 system [21,22]. These drugs provide Type-I interferon synthesis by raising serum retinoic acid levels to therapeutic levels and through retinoid signaling. These drugs include Remdesivir and Astemizol, which are approved by the FDA (US Food and Drug Administration). The presence of azole (benz-imidazole) ring in most of these drugs is also striking (Table 3).

**Table 3:**
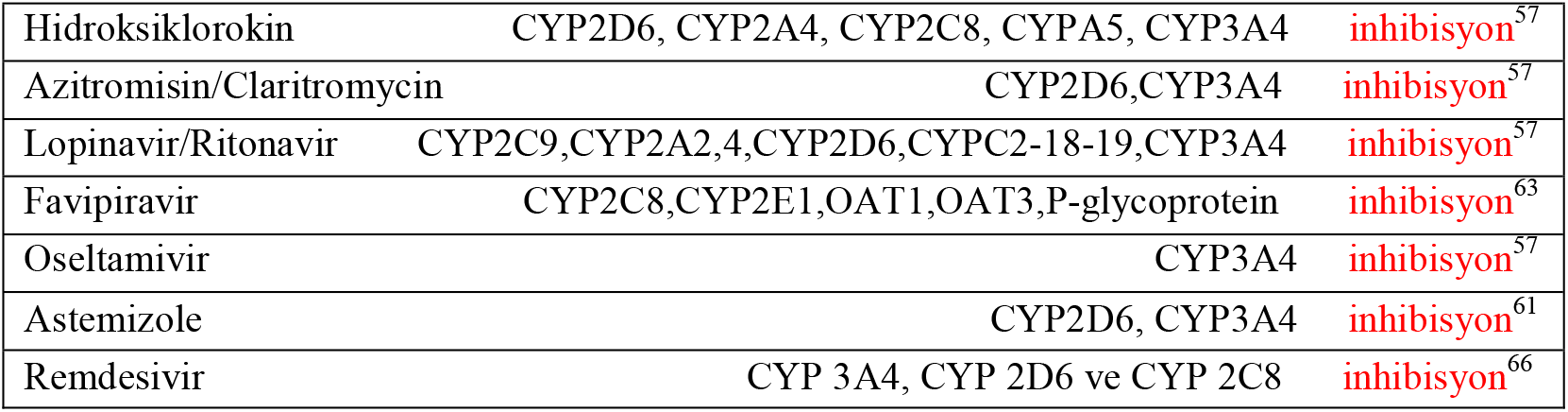
Inhibition/stimulation status of some drugs used in the treatment of COVID-19 on CYP450 enzymes.

The reason why the regulation mechanism of endogenous retinoic acids has not been noticed until now in COVID-19 may be the assumption that retinoic acid, an endogenous ligand, can always be present in the environment. However, the amount of retinoic acid in the human body is limited and is sufficient for approximately three months for a human [28]. Serum retinol levels drop only after the deficiency has progressed to severe levels because the liver contains large-scale stores of vitamin A. When serum retinol levels are found to be low, liver retinol stores will be largely depleted. Retinol and retinoic acids can be rapidly depleted due to reasons such as excessive viral load, high fever and catabolic destruction, especially with continuous and long-term RIG-I stimulation [28].

If some specific enzymes of the CYP450 system are inhibited, the metabolism of retinoic acids is inhibited, raising intracellular RA levels and therapeutic effects as in synthetic retinoids. For this purpose, agents that block the metabolism of retinoic acids (RAMBAs) have been developed [30]. RAMBAs have entered treatment applications in dermatological indications. This option is based on utilizing stored retinol esters in order to avoid the systemic and teratogenic side effects of retinoic acid derivatives [30]. Changes in CYP450 activity alter drug and endogenous metabolite concentrations in the body. This leads to an increase in endogenous metabolite levels and potential effects [21,30, 32].

Early treatment with drugs that inhibit CYP450 in COVID-19 increases serum retinoic acid levels by preventing retinoic acid metabolism through the liver. This effect manifests itself at the cellular level as gene expression and functional protein synthesis [28]. In this way, by providing Type-I interferon synthesis in the early period in COVID-19, the virus can be cleaned from the body without worsening the disease picture. With this early treatment, serum endogenous retinoic acid levels can be increased and cytokine storm and severe disease can be prevented by degradation inhibition in the ubiquitin proteosomal system (UPS) [28, 29].

### Vitamin A Use

Ten patients in the patient group received vitamin A and unsaturated fatty acids enterally and parenterally. Nine (90%) of them died. Of the 17 patients who did not take Vitamin A, 4 died (24%). There was a significant correlation between vitamin A use and dropout rates between these two groups (P <0.001). 90% of the patients who took vitamin A died. Although the difference in retinol between the group receiving vitamin A and the group not taking vitamin A was not significant, the mean retinol level was found to be higher in the group that received vitamin A. Eight of nine patients who took vitamin A and died had a comorbid disease. Therefore, it is impossible to tell whether the high output rate in this group is due to external Vitamin A use or a comorbid disease.

There is a paradoxical situation regarding the use of vitamin A under active inflammatory process or during active infection. Vitamin A metabolite all-trans retinoic acid (atRA) plays an important role in mucosal immune responses. atRA promotes effector T cell responses during infection or autoimmune diseases. Therefore, atRA plays a role in immune homeostasis at steady state, while activating pathogenic T cells under inflammation conditions [27, 28]. In addition, atRA modulates Foxp3 (+) regulatory T cell (Treg) and Th17 effector T cell differentiation. While it is used as an effective “mucosal adjuvant” in vaccines, it also appears to be necessary for the establishment of intestinal immune tolerance [14, 15, 16, 27, 33].

In addition, there are studies suggesting the use of high doses of vitamin A to regulate the immune system during infections. This understanding does not contradict the ability of vitamin A to boost immune responses and atRA’s ability to activate pathological T cells.^31^ These studies have shown that the effect of vitamin A is dose dependent. Accordingly, low-dose retinoic acids at physiological doses (1 nM) induce the release of inflammatory mediators by promoting conversion from T1 to T17, while pharmacological or high doses (10 nM or higher) retinoic acids inhibit T1 to T17 conversion and suppress inflammatory cytokine release and cause host infections [27]. It is argued that the immune system components that protect against it are activated. All these results suggest that RA may have a dose-differential effect on the differentiation of Th17 cells and Th1 cells [31]. There are views in the literature suggesting that retinoids and carotenoids may work in COVID-19 [34].

### Retinol and Type-I Interferon Gene Mutations in COVID-19

Recently, numerous studies on the defect in Type-I interferon synthesis in severe COVID-19 have been reflected in the literature [35, 36, 37, 38]. In these studies, the Type-I interferon synthesis defect has been attributed to mutations in some genes. However, subsequent studies have shown that these mutations occur only in 3-4% of severe COVID-19 patients, and these mutations cannot explain the Type-I interferon synthesis defect in COVID-19. In drug screening studies for COVID-19, Sumit Chandra and his colleagues found that retinol-dependent genes were not expressed in Vero-6 culture cells infected with SARS-CoV-2 [39].

The impaired retinoid signaling mechanism in COVID-19 causes Type-I interferon synthesis impairment. The healthy function of retinoid signaling depends on the presence of retinol and retinoic acid. Retinol and endogenous retinoic acids, which are rapidly depleted in the body, cause impairment of retinoid signaling. Disruption of retinoid signaling with the depletion of retinoic acids plays a triggering role in the pathogenesis of COVID-19 [40].

Retinoic acids and retinoid signaling play a decisive role in the development of severe clinical symptoms and signs in COVID-19. In viral infections, the Type-I IFN synthesis pathway used by TLR3 and RIG-I, which is a component of the innate immune system and where viral ssRNA ligands are first recognized, is of great importance. This pathway is the most active pathway in host defense until retinol and retinoic acids are exhausted [41, 42, 43]. The healthy functioning of this pathway depends on the presence of retinoic acid and Type-I interferon synthesis continues as long as the source of retinoic acid is available. Type-Interferon clears the pathogen from the body and provides a persistent immune response.

After the retinoic acids are exhausted, the RIG-I pathway is disabled and Type-I interferon synthesis stops. In the next process, the immune mechanism shifts to the NFKB pathway, which causes TNFα and excessive cytokine release via the TLR3, TLR7, TLR8, TLR9, MDA5 receptors and UPS pathway in neutrophil, monocyte, macrophage and dendritic cells of the adaptive immune system without retinoic acid. With the engagement of this mechanism, excessive TNFα and cytokine discharge occurs (cytokine storm) [40, 43, 44, 47]. The destructive excessive inflammatory response of the adaptive immune system that comes into play causes SIRS, ARDS and multiorgan injuries, aggravating the clinical picture. Adaptive immune system components consisting of monocyte, macrophage, neutrophil and dendritic cells are predominantly found in the lungs and intestines. The places where the inflammatory process bursts in COVID-19 are the structures associated with these two organs and the surrounding mucosal and submucosal immune system [45, 46].

COVID-19 patients have many entities and findings developed as a result of the depletion of retinol or endogenous retinoic acids. These include some laboratory and radiological imaging findings, clinical observations and symptoms [48]. These findings and symptoms that are seen in severe COVID-19 patients dramatically resemble the symptoms and findings seen in Vitamin A deficiency [49, 55]. Most of these symptoms and findings have been also reflected in the literature [49]. Some symptoms in COVID-19 patients such as particularly smell and taste disturbance, and symptoms involving the central nervous system, eye, cardiovascular system, digestion system, respiratory system, adrenal gland, kidney, testicle and especially immune system seem to be associated with the depletion of retinoic acids [50, 51, 52, 53]. Retinoic acid receptors and that are responsible for retinoic acid metabolism CYP450 enzymes are densely found in these systems and organs specified above [52,53,54]. The changes in cellular and the humoral immune system in COVID-19 dramatically resemble the immune system depression seen in Vitamin A deficiency [56, 57]. Smell and taste disturbance seen in COVID-19 is also commonly seen in isolated Vitamin A and zinc deficiency [57].

Another important issue is the question of whether the vaccines currently in use will work. Time will give the best answer to this question. Although these vaccines have passed their phase studies and started to be used, it is unclear how long and how effective they will be. Such concerns already existed due to the frequent mutations of SARS-CoV-2 [58]. However, the emergence of new SARS-CoV-2 variants in Europe and Africa has further fueled concerns about vaccines. In addition, organized anti-vaccination movements started to emerge in Europe and America [59, 60, 61, 62, 63]. Although 40 years have passed since HIV / AIDS, an effective vaccine has not yet been developed. With the drugs developed over time, AIDS has turned into a viable chronic disease [64, 65]. While these facts stand in front of us, pushing drug studies to the background may turn out to be a heavy return. Therefore, while vaccine development and vaccination studies continue, drug replacement and drug development studies for COVID-19 should be carried out intensively without pushing them to the second plan.

## Conclusion

COVID-19 occurs mildly in some people without any symptoms; however, it occurs very seriously in some. We think that this clinical difference is due to the state of retinol storage in the liver. Malnutrition, comorbid disease, chronic lung and liver diseases, obesity, hepatosteatosis, chronic inflammation, febrile diseases and excessive antigenic stimulation cause depletion of retinol stores and weaken immune defense against SARS-CoV-2. Type-I interferon response of the host to SARS-CoV-2 is associated with retinol storage status and endogenous retinoic acid levels. This study found that serum retinol levels were reduced in patients with severe COVID-19, and even retinol was on the verge of depletion in some patients. This suggests that the state of retinol stores and retinol depletion in COVID-19 has an impact on the clinical course.

A study to be conducted in a homogeneous COVID-19 patient group will be a guide for candidate drugs that may be effective for Type-I interferon synthesis and new drug targets for COVID-19 patients. Such a study is also needed to develop an effective immune response to vaccines to be prepared for COVID-19 and adjuvant applications. The fact that our study was conducted with a small number of patients and in homogeneous patient groups reveals the necessity of supporting this study with well-planned new studies. Given the potential for many overlooked factors to affect retinol levels, prospective clinical studies with larger, more carefully selected case groups are needed. However, we would like to emphasize that well-planned controlled clinical studies are needed to include Vitamin A or retinoic acid derivatives in COVID-19 treatment protocols. This study also sheds light on the pathogenesis of COVID-19 and guides COVID-19 treatment and prophylaxis.

A well-preserved serum retinol depot indicates that levels of endogenous retinoic acids can be an effective strategy against SARS-CoV-2 in host defense, especially in therapeutic ranges. Therefore, we can say that vitamin A and carotenoids can be effective in COVID-19 prophylaxis. Based on the available data, we predict that enrichment of plasma retinol storage can strengthen both the innate and adaptive immune systems in humans. Also, due to their potent immunomodulatory activities, retinol and retinoic acid derivatives, as well as some specific CYP450 inhibitors (RAMBAs), can act as prophylactic and therapeutic agents for COVID-19. Further epidemiological studies will enable the determination of vitamin A deficiency at the community level, feed the community with foods rich in retinoids and carotenoids, ensure the use of prophylactic retinol supplements when necessary, and thus protect the community from many viral infections, especially COVID-19.

## Data Availability

The sources referenced in the study are cited appropriately in the text and in the series of sources.
This article can be used provided that the authors are properly cited. Article content cannot be used for commercial purposes without the consent of the responsible author.

## Acknowledgments

I would like to thank Prof. Dr. Murat Kızıl and Prof. Dr. Mustafa Kemal Çelen who encouraged me for this work and did not withhold his support during the writing phase.

## Disclosure Statement

The authors are not a party of any affiliations, memberships, funding, or financial holdings that might be perceived as affecting the objectivity of this review.

## The Basis of the Study and the Decision of the Ethics Committee

This study was conducted with the approval of the ethics committee and the approval of Diyarbakır Gazi Yaşargil Training and Research Hospital, University of Health Sciences, and the approval of the Ministry of Health dated 03.06.2020 and numbered T22_10_40.xml. All protocols were implemented within the recommendations of the local ethics committee. All subjects were included in the study following the protocols approved by the local ethics committee. When the patients were hospitalized, they were informed about the study and were included in the study after obtaining their consent for signature.

## The Contents of the Formulas and TPN Administered to the Patients

The contents of the formulas and TPN administered to the patients were given below: More than one type of TPN and formulas were administered to some patients.

1. Resource® Diabet Nestlé Health Science. Administered to 3 individuals. 24 pcs, 10 pcs and 10 pcs. Protein: 28% E; 14 g/200 ml (Casein), Fat: 24% E; 5.4 g/200 ml (saturated, single-poly unsaturated fat), Carbohydrate: 44% E; 21.8 g/200 ml, Fiber: 4% E; A vial contains 4 g of soluble fiber. (Partially hydrolyzed guar gum).
2. Nutrivigor® Abbott. e220 ml 1.5 kcal/ml. 1.5 g CaHMB 330 kcal. Administered to 2 individuals, 10 pcs and 29 pcs. Contains 20g Protein, 500 IU Vitamin D.
3. Omegaven® 10% 100 ml infusion emulsion. Fresenius container. 3 pcs were used in 1 individual. 100 ml contains; 10.0 g fish oil refined at high temperature: 1.25-2.82 g Eicosapentaenoic Acid (EPA), 1.44-3.09 g Docosahexaenoic acid (DHA), 0.015-0.0296 g dl-a-Tocopherol, 2.5 g Glycerol, 1.2 g Purified egg phospholipids, 0-0.002 g Sodium Hydroxide (pH regulator), 0.003 g Sodium oleate (emulsifier).
4. Oligoclinomel N7-1000 E electrolyte amino acid solution, glucose solution, lipid emulsion.1500 ml three-chamber bag. 7 pcs were used in 1 individual. 100 ml contains refined olive oil (80%) + Refined soya oil (20%): 4 g, essential amino acids and Sodium acetate 3EE0: 0.245 g, Sodium glycerophosphate SEEO: 0.214 g, Potassium chloride: 0.179 g, Magnesium chloride 6H2O: 0.045 g, Glucose (17.6 g glucose monohydrate):16 g, Calcium chloride 2H2O: 0.030 g.
5. Novasource® GI Control 500 ml. Nestle. 8 pcs were used in 1 individual. Protein 15% E; 20.5 g/500 ml (milk protein), Carbohydrate 53% E; 72.5 g/500 ml, Fat 29% E; 17.5 g/500 ml. (saturated, single-poly unsaturated fat, MCT, omega-3).
6. Fortimel Energy. 200 ML. Nutricia. 28 pcs were used in 1 individual. 100 gram contains; 5.80 g Fat, 0.700 g Saturated Fat, 3.400 g Single Saturated Fat, 1.700 g Polyunsaturated Fat, 18.50 g Carbohydrates, 6.80 g Sugar, 5.80 g Protein
7. Resource® Energy 200ml. Nestlé Health Science. 38, 53 and 65 pcs were used in 3 individuals. Protein: 15% E-11.2 g/200 ml (Casein), Carbohydrate: 55% E-42 g/200 ml (Maltodextrin, sucrose), Fat: 30% E - 10 g/200 ml (saturated, single-poly unsaturated fat acids).
8. Cernevit im / iv Injectable / Lyophilized Powder. Baxter Healthcare Corporation. 3 people were used. There are 1, 4 and 5 pieces. In each vial: Vitamin A (as Retinol palmitate) 3500 IU, Vitamin D3 (Cholecalciferol) 220 IU, Vitamin E 11.2 IU, Vitamin C 125 mg, B Complex vitamins.

## Notes

### Competing Interest Statement

The authors have declared no competing interest.

### Funding Statement

No funding or similar incentives were received for any phase of the study.

### Author Declarations

This study was conducted with the approval of the ethics committee and the approval of Diyarbakır Gazi Yasargil Training and Research Hospital, University of Health Sciences, and the approval of the Ministry of Health dated 03.06.2020 and numbered T22_10_40.xml. All protocols were implemented within the recommendations of the local ethics committee. All subjects were included in the study following the protocols approved by the local ethics committee. When the patients were hospitalized, they were informed about the study and were included in the study after obtaining their consent for signature.

